# Relationship between blood pressure and the risk of acute myocardial infarction in Chinese adults: a prospective study

**DOI:** 10.1101/2023.03.03.23286788

**Authors:** Xin Zhang, Xiaoyong He, Fan Mao, Run Zhang, Xiaoqing You, Jianhong Li

**Affiliations:** National Center for Chronic and Noncommunicable Disease Control and Prevention, Chinese Center for Diseases Control and Prevention, Beijing 100050, China

**Keywords:** Hypertension, Acute myocardial infarction, Prospective study

## Abstract

**Objective:** To explore the relationship between blood pressure and the risk of acute myocardial infarction (AMI) in Chinese adults.

**Methods:** This prospective cohort study included 26,794 participants which were recruited from 60 surveillance sites in 2010 China Chronic Disease Risk Factor Surveillance and followed-up survey was conducted from 2016 to 2017. Data were collected by questionnaires, physical measurements and laboratory tests. Blood pressure was grouped by systolic blood pressure and diastolic blood pressure, and cox proportional hazards regression models were used to analyze the relationship between blood pressure and risk of AMI.

**Results:** A total of 256 AMI events occurred among 160,386.38 person-years follow-up and the incidence of AMI was 159.6/100,000 person-years. After adjusting for age, sex, income, education, marital status, physical activity, drinking, abdominal obesity, BMI, dyslipidemia and diabetes, compared with normotension group, the risk of AMI onset was increased by 61% (*HR*=1.61, 95% *CI*: 1.17∼2.21), 60% (*HR*=1.60, 95% *CI*: 1.17∼2.20), 84% (*HR*=1.84, 95% *CI*: 1.26∼2.68) and 120% (*HR=*2.20, 95% *CI*: 1.43∼3.38) in systolic diastolic hypertension(SDH), hypertension grade 1(mild), hypertension grade 2(moderate), hypertension grade 3(severe), respectively. In addition, after adjusting for relevant factors, the relationship between hypertension and the risk of AMI may be influenced by abdominal obesity, diabetes and lack of physical activity.

**Conclusions:** The incidence of AMI appears to be considerable among Chinese adults and hypertension is an independent risk factor for its onset. The risk of AMI is higher in participants with more severe hypertension.

---

Acute myocardial infarction (AMI), the myocardial necrosis caused by unstable ischemic syndrome, is the most serious manifestations of coronary heart disease. In practice, the diagnosis and evaluation of this disease are based on clinical assessment, electrocardiographic (ECG), biochemical tests, invasive and non-invasive imaging as well as pathological assessment^[1, 2]^. The onset of AMI causes serious consequence and relatively poor prognosis^[3]^. *Report on Cardiovascular Health and Diseases Burden in China (2020)* indicated that the mortality and morbidity of AMI showed an upward trend in recent years and further increased the economic burden to our society. Since 2004, the average growth rate of hospitalization expenses was 26.9% and the recurrence rate was higher at 35.7%. Moreover, case fatality rate among patients had a recurrent AMI (32.1%) was higher than the one-year case fatality among AMI patients (28%)^[4]^. Therefore, reducing the onset of AMI is considered an urgent task.

Numerous studies demonstrated that hypertension is a risk factor for the incidence of AMI^[5-7]^. However, studies on the relationship between hypertension and AMI are limited and most of those are clinical studies which efforts have been devoted to H-type hypertension or the relationship between pulse pressure and AMI^[8-11]^.Most previous epidemiological studies in this field have only focused on hypertension as one of the major risk factors for AMI or their correlation rather than comprehensively study the relationship between AMI and subtypes and severity of hypertension. Thus, this study aimed to fill this gap. It focused specifically on hypertension and grouped it into different subtypes and severity defined by systolic blood pressure (SBP) and diastolic blood pressure (DBP) to analyze the impact on the incidence of AMI. It was also hoped that this study would provide data support for our government to formulate corresponding prevention and control policies of AMI and put forward targeted suggestions for people with varying degrees of hypertension.

## 1. Participants and Methods

### 1.1 Participants

According to the 2010 China Chronic Disease Risk Factor Surveillance (CCDRFS), 60 surveillance sites were selected from 11 of the 31 provinces (Hebei, Jilin, Heilongjiang, Zhejiang, Jiangsu, Jiangxi, Henan, Hunan, Sichuan, Guizhou and Shanxi), which distributed in eastern, central and western and ranged from urban to rural regions. Excluding 157 participants with baseline AMI and 108 participants under 18years old, a total of 36,426 participants were included at baseline survey. A follow-up survey was conducted from 2016 to 2017 and the final follow-up survey was 27,497 participants (including 597 deaths and 56 among them due to AMI). Excluding 541 deaths due to other diseases and 162 participants with unknown information of body mass index (BMI) and waist circumference (WC), 26,794 participants were included in the analysis. This study passed the review of the Ethics Review Committee of National Center for Chronic and Noncommunicable Disease Control and Prevention, Chinese Center for Diseases Control and Prevention, (approval number 201524B), and all the participants signed the informed consent form.

### 1.2 Survey content

#### 1.2.1 Baseline survey

The baseline survey consisted of questionnaires through face-to-face survey, physical measurements, and laboratory tests. The questionnaire included basic personal information, lifestyle (diet, smoking and drinking, etc.), occurrence and diagnosis of chronic diseases and surveyed by trained and qualified investigators. Physical measurements included height, weight, waistline and blood pressure. The laboratory test requires each participant to collect 3 tubes of venous blood 5∼6ml after fasting for 10 to 12 hours, and another tube of venous blood 1ml after taking 75g glucose for 2 hours. The indexes included fasting plasma glucose, 2-hour plasma glucose after glucose load, glycosylated hemoglobin and blood lipid.

#### 1.2.2 Follow-up survey

The follow-up survey consisted of household and individual questionnaires. The individual questionnaire included basic personal information (age, sex, and marital status, etc.) and the onset and death of chronic diseases, etc. The household questionnaire included diet habits and economic status. The outcome of this study is the first occurrence of AMI (I21, according to ICD-10 classification) and the incidence information of AMI was obtained by questionnaire self-report. For identifying AMI case, participants were asked “whether AMI had ever been diagnosed by a physician since 2010” and whether that was the first onset during the in-person interview. Furthermore, diagnostic information was collected at the same time (including the basis of clinical diagnosis, imaging tests, time of diagnosis, diagnosis unit), and medical records and images were also reviewed. If the survey site has a Cardiovascular Event Reporting System, the questionnaire data was also be checked through the system. For participants who were lost to follow-up or reported dead during the survey were verified by obtaining information from National Cause of Death registry, found out details and complemented relevant information. If participants were unable to participate in the survey due to physical or absence, a proxy participant (usually were family members familiar with the situation) were identified to complete the survey for them.

### 1.3 Risk factors

(1) Dyslipidemia: according to *2016 Chinese guideline for the management of dyslipidemia in adults*^[12]^, dyslipidemia was defined as triglyceride (TG) ⩾2.26 mmol/L or total cholesterol (TC) ⩾ 6.22 mmol/L or low density lipoprotein cholesterol (LDL-C) ⩾4.14 mmol/L or high density lipoprotein cholesterol (HDL-C) <1.04 mmol/L. (2) Abdominal obesity: the WC of men ⩾85 cm or the WC of women ⩾80 cm. (3) Insufficient intake of fruits and vegetables: according to the World Health Organization recommendation, the average daily intake of less than 400g of fruits and vegetables was defined as insufficient intake. (4) Excessive intake of red meat: the average daily intake above 100g was defined as excessive intake according to the recommendation of the World Cancer Research Fund. (5) Smoking: participants who were still smoking at the time of survey were defined as smokers. (6) Drinking: participants who reported drinking any purchased or homemade alcoholic beverages (including liquor, beer, red wine, wine, fruit wine, yellow rice wine and highland barley wine, etc.) during the past 12 months were defined as drinker. (7) Lack of physical activity: According to the physical activity metabolic equivalent (MET) value of the physical activity outline put forward by Ainsworth^[13]^, physical activity was divided into high, medium, and low intensity, and the corresponding MET values were 8.0, 4.0, and 3.3, respectively. The amount of physical activity per week was equal to the sum of the MET of various intensities multiplied by the weekly activity time (min/week). Physical activity of less than 600 MET-min/week was defined as physical inactivity.

### 1.4 Index definition and grouping

(1) Blood pressure grouping: according to *Chinese guidelines for the management of hypertension (2018 revised edition)*^[14]^, blood pressure can be categorized into normotension (SBP< 140mmHg(1mmHg = 0.133kPa) and DBP< 90mmHg) and hypertension(SBP ≥ 140mmHg and/or DBP ≥ 90mmHg). Generally, hypertension can be classified into three subtypes by level of SBP and DBP: 1) Isolated systolic hypertension (ISH): SBP≥ 140mmHg and DBP< 90mmHg; 2) Isolated diastolic hypertension (IDH): SBP< 140mmHg and DBP≥ 90 mmHg; 3) Systolic diastolic hypertension (SDH): SBP≥ 140mmHg and DBP≥ 90mmHg. In addition, hypertension can be divided into three grades: 1) Hypertension grade 1 (mild): 140 ≤ SBP < 159mmHg and/or 90 ≤ DBP < 99mmHg; 2) Hypertension grade 2 (moderate): 160≤ SBP< 179mmHg and/or 100≤ DBP< 109mmHg; 3) Hypertension grade 3 (severe): SBP≥ 180mmHg and/or DBP≥ 110mmHg.

(2) Obesity grouping: obesity was defined based on BMI according to the *Guidelines for Prevention and Control of Overweight and Obesity in Chinese Adults*^[15]^. BMI was calculated by dividing weight in kilograms by height in meters squared. It can be divided into four groups: underweight(BMI<18.5 kg/m^2^), normal weight(18.5 ⩽ BMI⩽ 23.9 kg/m^2^), overweight(24⩽ BMI⩽ 27.9 kg/m^2^) and obesity(BMI⩾ 28.0 kg/m^2^).

### 1.5 Statistical analysis

We used SAS9.4 software for data collation and analysis. For continuous variables, mean standard deviation was used to describe normal distribution or approximate normal distribution, while those who did not accord with normal distribution were described by *M* (P_25_, P_75_). For the classified data, the number of cases and constituent ratio n (%) were used to describe the classification data. F-test, Chi-square test and Wilcoxon rank sum test or Kruskal-Wallis rank sum test were used for inter-group comparison. Cox proportional hazards model was used to calculate hazard ratios (*HR*) and 95% confidence intervals (95% *CI*) for the relationship between blood pressure and AMI. The statistical significance level was set at *P*< 0.05.

## 2. Results

### 2.1 Baseline Characteristics

We analyzed data from 26,794 participants. Their median age was 47.7 years with a male/female ratio of 100: 123 and most were married (84.25%). Participants who were aged between 18 and 45 accounted for the largest proportion (41.94%) and most educational level of participants was primary school or less than primary school (45.81%). The prevalence of overweight, obesity, abdominal obesity, diabetes, dyslipidemia was 33.21%、13.49%、42.39%、6.67% and 51.77%, respectively. In addition, among this study participants, 27.55% were smokers of which 6886(93.27%) were male; 36.39% were drinkers of which 7323(75.11%) were male; 55.32% with daily intake of fruits and vegetables less then 400 g; 12.60% consumed red meat more than 100 g per day and 18.07% were lack of physical activity(Table)

### 2.2 Relationship between blood pressure and the risk of AMI

A total of 256 AMI events occurred and the incidence of AMI was 159.6/100,000 person-years during 160386.38 person-years follow-up. After adjusting for age, gender, income, education, marital status, physical activity, drinking, abdominal obesity, BMI, dyslipidemia and diabetes, compared with normotension group, the risk of AMI onset was increased by 61% (*HR*=1.61, 95% *CI*: 1.17∼2.21), 60% (*HR*= 1.60, 95% *CI*: 1.17∼2.20), 84% (*HR*=1.84, 95% *CI*: 1.26∼2.68) and 120% (*HR*= 2.20, 95% *CI*: 1.43∼3.38) in SDH, hypertension grade 1(mild), hypertension grade 2 (moderate), hypertension grade 3 (severe) group, respectively. However, ISH group and IDH group made no significant difference to the risk of AMI (*P*>0.05)(table2).

**Table 1.** Baseline characteristic of participants.

| Variable | Man<br>(n=12,019) | Woman<br>(n=14,775) | Total<br>(n=26,794) |
| --- | --- | --- | --- |
| Age (year), $\bar{x} \pm s$ | 47.9±13.9 | 47.6±13.5 | 47.7±13.7 |
| Age grouping, n (%) |  |  |  |
| 18~45 | 5,003(41.63) | 6,235(42.20) | 11,238(41.94) |
| 45~60 | 4,443(36.97) | 5,661(38.31) | 10,104(37.71) |
| $\geq 60$ | 2,573(21.41) | 2,879(19.49) | 5,452(20.35) |
| Education, n (%) |  |  |  |
| Illiterate | 4,499(37.43) | 7,774(52.62) | 12,273(45.80) |
| Primary school | 4,640(38.61) | 4,272(28.91) | 8,912(33.26) |
| Junior school | 1,985(16.52) | 1,814(12.28) | 3,799(14.18) |
| Senior high school and above | 895(7.45) | 915(6.19) | 1,810(6.76) |
| Marital status, n (%)<sup>a</sup> |  |  |  |
| Unmarried | 968(8.08) | 544(3.69) | 1,512(5.65) |
| Married / cohabiting | 10,094(84.24) | 12,480(84.61) | 22,574(84.45) |
| Widowed / divorced / separated | 920(7.68) | 1,726(11.70) | 2,646(9.90) |
| Total household income, n (%) |  |  |  |
| <10000 | 3,816(31.75) | 4,895(33.13) | 8,711(32.51) |
| 10000~20000 | 2,685(22.34) | 3,434(23.24) | 6,119(22.84) |
| 20000~30000 | 1,988(16.54) | 2,320(15.70) | 4,308(16.08) |
| $\geq 30000$ | 3,530(29.37) | 4,126(27.93) | 7,656(28.57) |
| BMI (kg/m<sup>2</sup>), n (%) |  |  |  |
| <18.5 | 365(3.04) | 489(3.31) | 854(3.19) |
| 18.5~24 | 6,254(52.03) | 7,173(48.55) | 13,427(50.11) |
| 24~28 | 3,965(32.99) | 4,934(33.39) | 8,899(33.21) |
| $\geq 28$ | 1,435(11.94) | 2,179(14.75) | 3,614(13.49) |
| Blood pressure grouping, n (%) |  |  |  |
| normotension | 7,407(61.63) | 9,451(63.97) | 16,858(62.92) |
| ISH | 1,758(14.63) | 2,143(14.50) | 3,901(14.56) |
| IDH | 529(4.40) | 493(3.34) | 1,022(3.81) |
| SDH | 2,325(19.34) | 2,688(18.19) | 5,013(18.71) |
| hypertension grade1 | 2,862(23.81) | 3,051(20.65) | 5,913(22.07) |
| hypertension grade 2 | 1,222(10.17) | 1,477(10.00) | 2,699(10.07) |
| hypertension grade 3 | 528(4.39) | 796(5.39) | 1,324(4.94) |
| Urban, n (%) | 4,519(37.60) | 5,986(40.51) | 10,505(39.21) |
| Somking, n (%) | 6,886(57.29) | 497(3.36) | 7,383(27.55) |
| Drinking, n (%) | 7,323(60.93) | 2,427(16.43) | 9,750(36.39) |
| Abdominal obesity, n (%) | 4,627(38.50) | 6,732(45.56) | 11,359(42.39) |
| Diabetes, n (%) | 709(5.90) | 1,078(7.30) | 1,787(6.67) |
| Dyslipidemia, n (%) | 6,680(55.58) | 7,192(48.68) | 13,872(51.77) |
| Insufficient intake of fruits and vegetables, n (%) | 6,772(56.34) | 8,050(54.48) | 14,822(55.32) |
| Excessive intake of red meat, n (%) | 1,972(16.41) | 1,403(9.50) | 3,375(12.60) |
| Lack of physical activities, n (%) | 2,475(20.59) | 2,368(16.03) | 4,843(18.07) |
| SBP (mmHg), $\bar{x} \pm s$ | 135.2±20.1 | 133.5±23.2 | 134.2±21.8 |
| DBP (mmHg), $\bar{x} \pm s$ | 82.2±11.5 | 81.2±11.7 | 81.7±11.6 |
| TG (mmol/L), $\bar{x} \pm s$ | 1.5±1.6 | 1.4±1.2 | 1.5±1.4 |
| TC (mmol/L), $\bar{x} \pm s$ | 4.1±1.1 | 4.1±1.1 | 4.1±1.1 |
| HDL-C (mmol/L), $\bar{x} \pm s$ | 1.1±0.3 | 1.1±0.3 | 1.1±0.3 |
| LDL-C (mmol/L), $\bar{x} \pm s$ | 2.3±0.8 | 2.3±0.8 | 2.3±0.8 |
| WC (cm), $\bar{x} \pm s$ | 82.7±10.2 | 79.8±9.9 | 81.1±10.1 |
| FPG (mmol/L), $\bar{x} \pm s$ | 5.6±1.4 | 5.5±1.3 | 5.5±1.4 |
| 2-hour plasma glucose (mmol/L), $\bar{x} \pm s$ | 6.1±2.6 | 6.4±2.5 | 6.3±2.5 |

Note. BMI, body mass index; ISH, isolated systolic hypertension; IDH, isolated diastolic hypertension; SDH, systolic diastolic hypertension; SBP, systolic blood pressure; DBP, diastolic blood pressure; TG, triglyceride; TC, total cholesterol; HDL-C, high density lipoprotein cholesterol; LDL-C, low density lipoprotein cholesterol; WC, waist circumference; FPG, fasting plasma glucose; <sup>a</sup>lack of 62cases

**Table 2.** Relationship between blood pressure and the risk of AMI.

|  | Number of events | Follow-up year | Incidence<br>(no./1,000 person-years) | HR(95%CI) |  |  |
| --- | --- | --- | --- | --- | --- | --- |
|  |  |  |  | Model 1 | Model 2 | Model 3 |
| Blood pressure grouping 1 |  |  |  |  |  |  |
| Normotension (n=17,491) | 101 | 10,1011.7 | 1.0 | 1.00 | 1.00 | 1.00 |
| ISH(n=4,027) | 62 | 23,300.9 | 2.7 | 1.53(1.09~2.14) | 1.49(1.06~2.11) | 1.30(0.91~1.84) |
| IDH(n=1,066) | 7 | 6,125.6 | 1.1 | 1.37(0.63~2.95) | 1.35(0.63~2.93) | 1.12(0.52~2.44) |
| SDH(n=5,170) | 86 | 29,948.1 | 2.9 | 2.08(1.53~2.81) | 2.00(1.47~2.72) | 1.61(1.17~2.21) |
| Blood pressure grouping 2 |  |  |  |  |  |  |
| normotension(n=17,491) | 101 | 101,011.7 | 1.0 | 1.00 | 1.00 | 1.00 |
| hypertension grade1(n=6,124) | 76 | 35,368.0 | 2.1 | 1.56(1.15~2.13) | 1.56(1.14~2.14) | 1.60(1.17~2.20) |
| hypertension grade 2(n=2,781) | 45 | 16,101.3 | 2.8 | 1.88(1.30~2.73) | 1.78(1.23~2.59) | 1.84(1.26~2.68) |
| hypertension grade 3(n=1,358) | 34 | 7,905.4 | 4.3 | 2.41(1.60~3.62) | 2.27(1.50~3.43) | 2.20(1.43~3.38) |

Note. ISH, isolated systolic hypertension; IDH, isolated diastolic hypertension; SDH, systolic diastolic hypertension; Model1 adjusts age and gender; Model2 adjusts income, education, marital status, physical activity and alcohol drinking on the basis of model1; Model3 adjusts abdominal obesity, BMI, dyslipidemia and diabetes on the basis of model2.

### 2.3 Subgroup analysis of the association between blood pressure and the risk of AMI

After adjusting the related influencing factors, no effect modification was found on the association between blood pressure and the risk of acute myocardial infarction in each variable subgroup of SDH outcome (all interactive P > 0.05). In the grading of hypertension, central obesity, diabetes and lack of physical activity had an effect on the risk of hypertension and AMI (all P < 0.05), but no interaction was found among the other subgroups (all P > 0.05) (Figure1).

**Figure 1.**
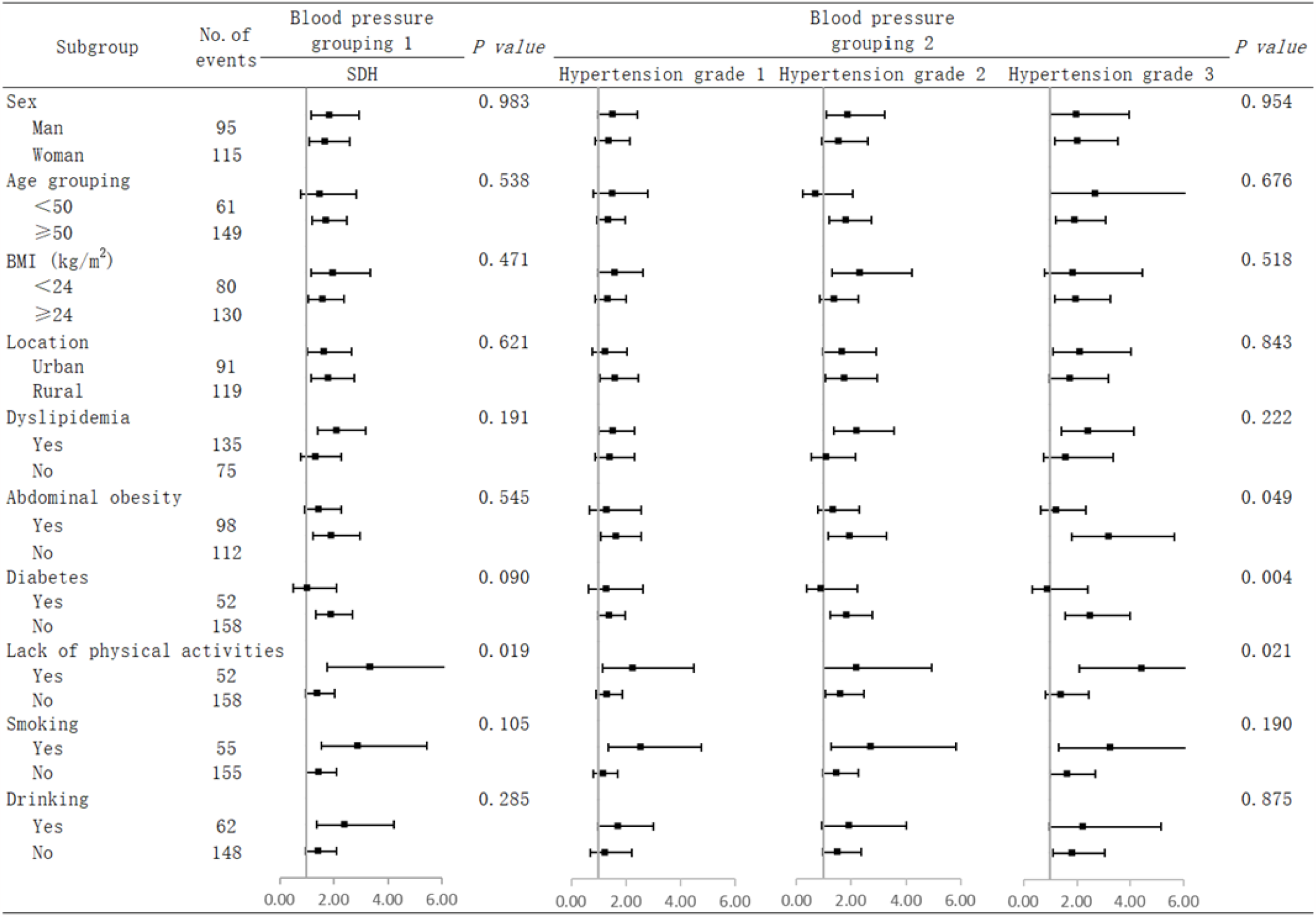
Subgroup analysis of the association between blood pressure and the risk of AMI. Note. Report the risk effect values of AMI in SDH, hypertension grade 1, hypertension grade 2 and hypertension grade 3 group with reference to normotension group; model adjusts age, gender, location, dyslipidemia, abdominal obesity, diabetes, lack of physical activity, smoking, drinking, excluding stratification factors.

## DISCUSSION

This study undertook a large population-based cohort study to explore the relationship between blood pressure and AMI in Chinese adults and studied incidence among respondents with and without specific underlying risk conditions, assessing the influence of these conditions on the risk of developing AMI in these population. As the main findings, its data indicates that the incidence of AMI in Chinese adults (159.6/100000 person-years) is considerable and hypertension is an independent risk factor of AMI.

Recent studies conducted in provincial administrative regions of China have demonstrated that the incidence of AMI is on the rise^[16-20]^. In Chongqing, the incidence and standardized incidence of AMI increased from 29.86/100,000 and 24.52/100,000 in 2012 to 52.67/100,000 and 39.56/100,000 in 2018, respectively^[19]^. Several studies found that the increasing trend of AMI incidence in China is associated with increased risk factors, such as, diabetes, aging population and cholesterol, and poor hypertension control^[5, 21]^. While compared with China, both age-standardized incidence and mortality of AMI in most Western countries have declined since the 1960s^[22-24]^. The explanation for this is that their awareness, treatment and control of three key risk factors (including hypertension, hypercholesterolemia and smoking) have improved^[25-27]^. However, as the process of urbanization and industrialization in China has accelerated in recent years, the lifestyle of residents has greatly changed and unhealthy lifestyles have been widely prevalent, leading to increased prevalence of hypertension, dyslipidemia, diabetes and obesity. In addition, a previous study also confirmed that, blood pressure elevation shares some common mechanisms with AMI, with pathophysiological links including endothelial dysfunction, autonomic nervous system dysregulation, impaired vascular reactivity, and genetic substrates^[28]^. These findings suggest that it is crucial to control the risk factors such as hypertension to effectively prevent and reduce the incidence of AMI in China. So good long-term blood pressure control should be provided for hypertensive patients.

After adjusting for age, gender, economic status, education, marital status, physical activity, alcohol drinking, abdominal obesity, BMI, dyslipidemia and diabetes, the risk of AMI onset was increased by 61% in the SDH group compared to normotension, while ISH and IDH were not related to the risk of AMI. However, some studies are different from the result of this study, which show that SBP is a risk factor for AMI, and larger studies are needed to analyze the relationship between SBP and risk of AMI. Studies have also shown that participants with SDH have the highest risk of any cardiovascular event among untreated hypertensive patients, while both ISH and IDH increase the risk of cardiovascular disease compared with normotensive participants^[29]^. A possible explanation for this different finding might be that outcomes included not only AMI but also stroke and death. Because of this inconsistency, additional studies exploring the relationship between ISH and IDH and AMI are needed to confirm the results of this study.

Another important finding about different hypertension grade of this study is that compared with normotension group, the risk of AMI onset was increased by 60%, 84% and 120% in hypertension grade 1, hypertension grade 2 and hypertension grade 3 group, respectively. This indicates that the more severe the hypertension, the higher the risk of AMI onset. To reduce AMI risk, emphasis should be placed on hypertension prevention and control, especially moderate and severe hypertension. Thus, in daily life, hypertensive patients should reduce sodium intake and perform appropriate physical activities to control their weight for long-term blood pressure control.

Subgroup analyses revealed that, after adjusting the relevant influencing factors, among groups classified by hypertension grade, relationship between hypertension and AMI was found affected by abdominal obesity, diabetes and lack of physical activity. This finding is consistent with previous studies which mentioned that increased the incidence of AMI is associated with increased risk factors such as diabetes and cholesterol^[5,21]^. Therefore, more attention should be paid to blood pressure control in people with abdominal obesity, diabetes and physical inactivity to more effectively prevent the occurrence of AMI.

This study has the strength of a prospective cohort study. Additionally, unique strength of this study is that it simultaneously analyzed the relationship between AMI and the classification and grade of hypertension, which previous studies rarely have done. And its relatively large sample size of which participants distributed across eleven provinces of the country has strength as a nationally representative study. However, several limitations to this study need to be acknowledged. Firstly, as a prospective study, there is a certain lost population which may lead to underestimation or overestimation of the incidence of AMI, even though the loss-to-follow-up rate is within the standard range (<30%). Secondly, since there are so many factors affecting AMI, that the baseline survey did not collect all the possible influencing factors, and some unmeasured indicators (such as mental factors) were not included in the model adjustment, which may lead to residual confounding. These limitations provide a direction for the study of the incidence and influencing factors of AMI in the future.

In conclusion, the results of this study show that the incidence of AMI is 159.6/100,000 person-years in Chinese adults and hypertension is an independent risk factor for its onset. The evidence from this study also points out that more severe hypertension is associated with higher risk of AMI onset. Therefore, for patients with hypertension, more intensive hypertension controlling and more frequent daily blood pressure and AMI monitoring may be required to reduce the onset of AMI.

## Data Availability

Data are available and can be obtained by contacting the corresponding author.

## Declarations

### Ethics approval and consent to participate

This study had been passed the review of the Ethics Review Committee of the National Center for Chronic and Noncommunicable Disease Control and Prevention, Chinese Center for Disease Control and Prevention (approval number: 201524B). All methods had been performed in accordance with the Declaration of Helsinki. All the participants signed the informed consent form, and for the illiterate participants, the informed consent of their parents and/or legal guardians was obtained.

### Consent for publication

No applicable.

### Availability of data and materials

The datasets used and analyzed during the current study available from the corresponding author on reasonable request.

### Authors’ contributions

**Xin Zhang and Xiaoyong He:** Conceptualization, Software, Formal analysis,Writing-Original Draft **Fan Mao, Run Zhang and Xiaoqing You:** Investigation, Resources, Data Curation **Jianhong Li:** Writing-Reviewing and Editing.

## Acknowledgements

No applicable.

